# Changes in transmission of Enterovirus D68 (EV-D68) in England inferred from seroprevalence data

**DOI:** 10.1101/2021.11.08.21265913

**Authors:** Margarita Pons-Salort, Ben Lambert, Everlyn Kamau, Richard Pebody, Heli Harvala, Peter Simmonds, Nicholas C Grassly

**Affiliations:** MRC Centre for Global Infectious Disease Analysis, School of Public Health, Imperial College London, UK; Department of Computer Science, University of Oxford, UK; Nuffield Department of Medicine, University of Oxford, UK; Immunization Department, Public Health England, UK; Infection and Immunity, University College of London, UK

## Abstract

The factors leading to the global emergence of enterovirus D68 (EV-D68) in 2014 as a cause of acute flaccid myelitis (AFM) in children are unknown. To investigate potential changes in virus transmissibility or population susceptibility, we measured the seroprevalence of EV-D68-specific neutralising antibodies in serum samples collected in England in 2006, 2011 and 2017. Using catalytic mathematical models, we estimate an approximately two-fold increase in the basic reproduction number over the 10-year study period, coinciding with the emergence of clade B around 2009. Despite such increase in transmission, the virus was already widely circulating before the AFM outbreaks and the increase of infections by age cannot explain the observed number of AFM cases. Therefore, the acquisition of or an increase in neuropathogenicity would be additionally required to explain the emergence of outbreaks of AFM. Our results provide evidence that changes in enterovirus phenotypes cause major changes in disease epidemiology.

## Introduction

Interest in understanding the epidemiology and disease impact of enterovirus D68 (EV-D68) and other enteroviruses has increased in recent years. Contrary to most human enteroviruses, EV-D68 causes severe respiratory disease and is transmitted by the respiratory route, sharing properties with rhinoviruses (1). Although this virus was first isolated in 1962, for decades it was only reported from isolated cases or small case clusters of respiratory disease (2).

From 2009-2010 onwards however, an increasing number of outbreaks of EV-D68-associated severe respiratory illness (SRI) have been reported worldwide (3, 4). In 2014, the United States experienced the first big outbreak of respiratory disease linked to EV-D68, with >1,100 cases reported by the Centers for Disease Control and Prevention (5). In parallel with this outbreak, an unusual number of acute flaccid myelitis (AFM) cases (a newly recognized condition that includes the sudden onset of flaccid limb weakness (6)) was also reported, and similar AFM outbreaks subsequently occurred in 2016 and 2018 (7). Retrospectively, it now appears, an unusual spike of ‘polio-like’ cases reported in 2012 in California (8) was an early occurrence of what was subsequently defined as AFM. In the UK and elsewhere in Europe, AFM cases have also been reported in recent years associated with upsurges in EV-D68 detections (9-11). Evidence that EV-D68 is the main cause of these AFM outbreaks has been growing (7, 12), although the role of other enterovirus serotypes such as enterovirus A71 (EV-A71) has not been discounted (13). There is no effective treatment or vaccine for EV-D68 infection yet, and residual paralysis and neurological sequelae after AFM is common and lifelong.

The mechanisms that have led to the emergence of EV-D68 outbreaks since the late 2000s remain unknown. One hypothesis is that transmission has increased as a result of evolutionary selection for increased replication fitness, or through the appearance of immune escape-associated mutations that lead to the evasion of pre-existing population immunity. Another is that the virus has become more pathogenic, and, as a consequence, the number of symptomatic (and therefore, reported) infections has increased independently of its transmissibility (i.e. the virus already circulated in the past but went mostly undetected).

As for other enterovirus serotypes, many EV-D68 infections are asymptomatic or mild and self-limiting. In addition, enterovirus surveillance is passive in most countries, based on laboratory reporting for samples submitted by clinicians for testing. It is consequently difficult to determine the true incidence of infection or whether changes in EV-D68 circulation have occurred. A recent study based on data from the BioFire FilmArray Respiratory Panel (14) has shown biennial cycles of EV-D68 circulation in the US at the national level since 2014, coinciding with the years of AFM outbreaks (7). However, these data are limited before 2014 and respiratory samples or throat swabs are infrequently tested by enterovirus surveillance programmes in the US or elsewhere. Seroprevalence surveys therefore offer an attractive potential alternative opportunity to investigate patterns of exposure to EV-D68. Detection of EV-D68 antibodies with adequate sensitivity and specificity can indicate prior infection and can be analysed using mathematical models to infer trends in the incidence of infection over time, by age-group and location.

Here, we use data on the prevalence of neutralising antibodies against EV-D68 from opportunistically collected serum samples broadly representative of the general population in England in 2006, 2011 and 2017 to reconstruct long-term changes in EV-D68 transmission. Using a mathematical model-based framework, we estimate changes in the annual force of infection (FOI) and the basic reproduction number (*R*_0_) and reconstruct the estimated annual number of new infections in each age class.

## Results

Individuals are assigned an antibody titer as the highest antibody dilution (1:4, 1:8,…, 1:2048) preventing virus replication (i.e. showing neutralization) (15). For EV-D68, it is unknown which neutralising antibody titer (or seropositivity cut-off) is indicative of true past infection. A simple method to determine a seropositivity cut-off is based on fitting a mixture model to the individual titer distribution, in order to differentiate between two sub-populations (seronegatives and seropositives) (16). However, determining such a cut-off was not possible here, as for two of the three serosurveys and for all the data combined, the distributions did not show a bi-modal shape (Figure S1). We therefore present our modelling analysis for two different cut-offs: a first weak cut-off of 1:16, which has been previously used in the literature to define EV-D68 seropositivity (15, 17), and a more stringent cut-off of 1:64, which we show next that provides seroprevalence curves by age similar to an even more stringent cut-off of 1:128 (Figure 1).

**Figure 1.**
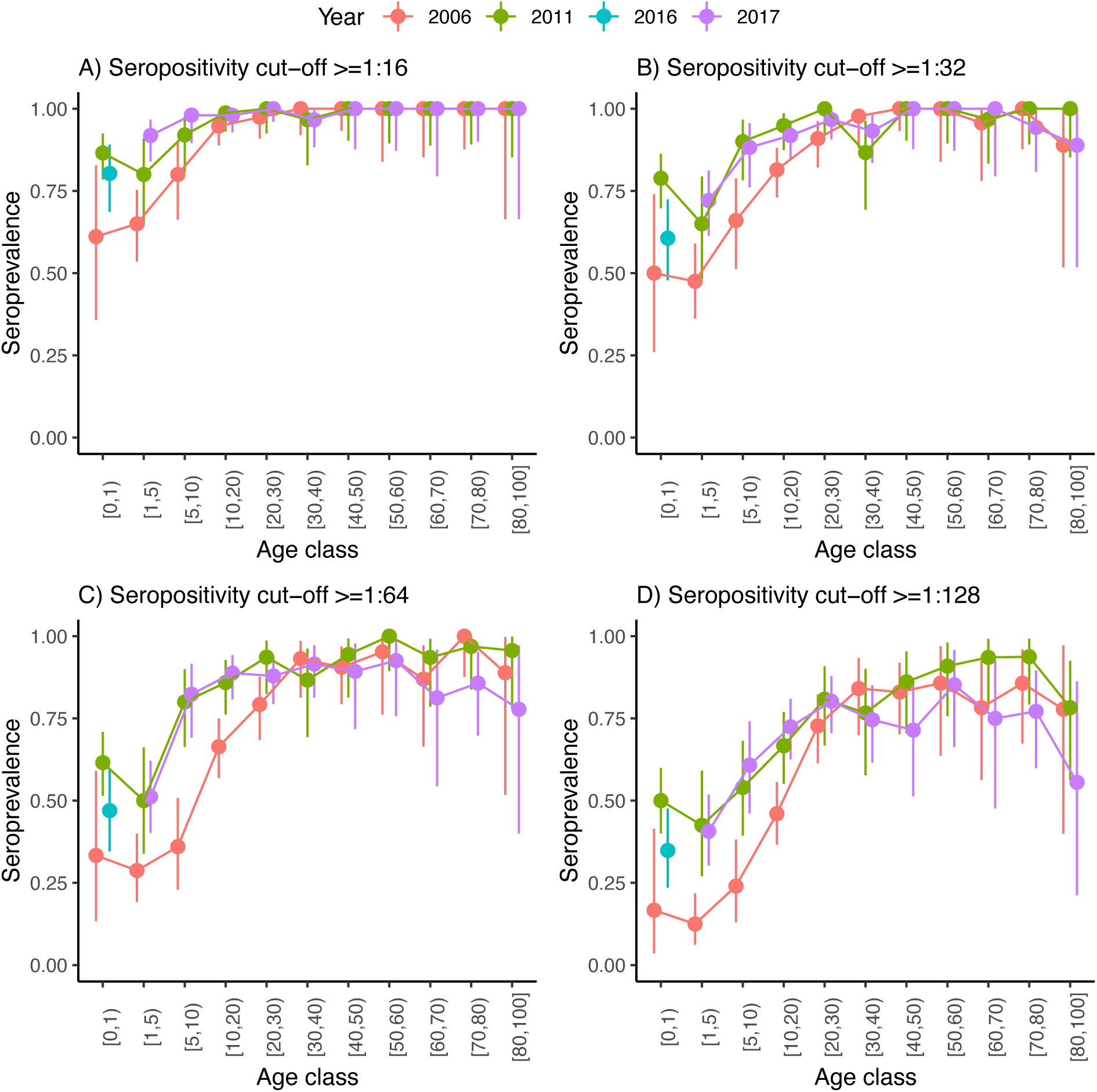
Seroprevalence by age in years. Seroprevalence by age class for the three serosurveys and for different cut-offs of neutralising antibody titre used to define positivity: (A) 1:16, (B) 1:32, (C) 1:64 and (D) 1:128. Bars are 95% binomial confidence intervals. Note that individuals in the [0-1) age class from the 2017 serosurvey were in fact sampled in 2016 and as such, are shown with a different colour. These individuals were not included in the analysis, as all individuals <1 yo were excluded to avoid the effect of maternal antibodies (Materials and Methods).

High rates of acquisition of EV-D68 infection could be inferred from seroprevalence frequencies in different age groups from the three serosurveys of samples collected in 2006, 2011 and 2017 (Figure 1). At each timepoint, irrespective of the cut-off antibody titre chosen to define seropositivity, seroprevalence slightly decreases from the 0 years-old (yo) to the 1-4 yo age classes, and then increases sharply with age until the 20-29 yo, when it reaches a plateau (Figure 1). As for many other viruses, higher values in the 0-yo age class are likely the result of the presence of transplacentally acquired maternal antibodies that subsequently decline over the following 6-12 months with lowest antibody titers and seroprevalence in the 6-12 month age class for EV-D68 (15). For a seropositivity cut-off of 1:16, the proportion seropositive at ages 1-4 yo ranged between 0.65 (95% CI 0.54 – 0.75) in 2006 and increased to 0.92 (95% CI 0.84 – 0.97) in 2017. For a more stringent cut-off of 1:64, the proportion seropositive in this age group decreased to 0.29 (95%CI 0.19 – 0.40) in 2006 and 0.51 (95% CI 0.40 – 0.62) in 2017. Age-stratified seroprevalence was generally lower in 2006 compared to 2011 and 2017, which suggests a decrease in the mean age of exposure through the study period, potentially consistent with increased transmission.

We first conducted exploratory analysis using two simple catalytic models that assumed the risk of infection (becoming seropositive) was independent of age and that the FOI was constant over time (Model 1), or that it was different and independent each year (Model 2) (Materials and Methods). As expected, Model 1 did not capture the different changes in seroprevalence by age among the three studies (Figure S2). Model 2 was able to capture these differences (Figure S3) but resulted in unrealistically high estimates of the annual probability of infection for the years the studies were conducted in order to reproduce the high seropositivity rates observed at age 1 (Figure S4).

Based on the results of the exploratory analyses (Models 1 and 2), we developed three further models (Models 3 to 5) with fewer free parameters than Model 2 but that allowed us to address the question of whether transmission had increased over time (before the first reported big outbreak of EV-D68 in the US in 2014). Model 3 uses a step-function to account for a single change in the FOI over time, and therefore consists of two constant FOI periods separated by an instantaneous step at a to-be-estimated timepoint (year), *T*. Model 4 assumes that the FOI changes over time following a random walk. For Models 3 and 4, we further assume that individuals of 1 year of age (i.e. 12-23 months old) have a higher risk of infection than subsequent age classes, which is modelled as the product of the FOI in a given year multiplied by a coefficient **α** that is estimated. The last model, Model 5, expands Model 4, and further assumes that the risk of infection decreases exponentially with age, with a coefficient **β** that is also estimated. See details of the models in the Materials and Methods section.

Models 3 to 5 provided a good fit to the data (Figures 3, S5 and S6) and all estimated an increase in transmission over time independent of the seropositivity cut-off that was used, as shown by the estimated annual probability of infection in Figure 2. The step-function model (Model 3) estimates an approximately 2-fold increase in the annual probability of infection using both seropositivity cut-offs (median of 0.11 in the first period vs. 0.21 in the second one for the 1:16 cut-off, and 0.06 vs. 0.15 for the 1:64 cut-off) (Figures 2A and 2B). When using the weak cut-off (1:16), the change is estimated to occur around 2007 (95% CrI 2005 – 2011), whereas for the more stringent cut-off, the model estimated a slightly earlier change in the FOI, around 2006 (95% CrI 2003 – 2008) (Figure S7). The models based on a random walk (Models 4 and 5) estimated a smooth and slow increase in the annual probability of infection that started in the early 2000s and continued until around 2010, when using a seropositivity cut-off of 1:64 (Figures 2D and 2F), or continued until the end of the study period, in 2017, for a cut-off of 1:16 (Figures 2C and 2E).

**Figure 2.**
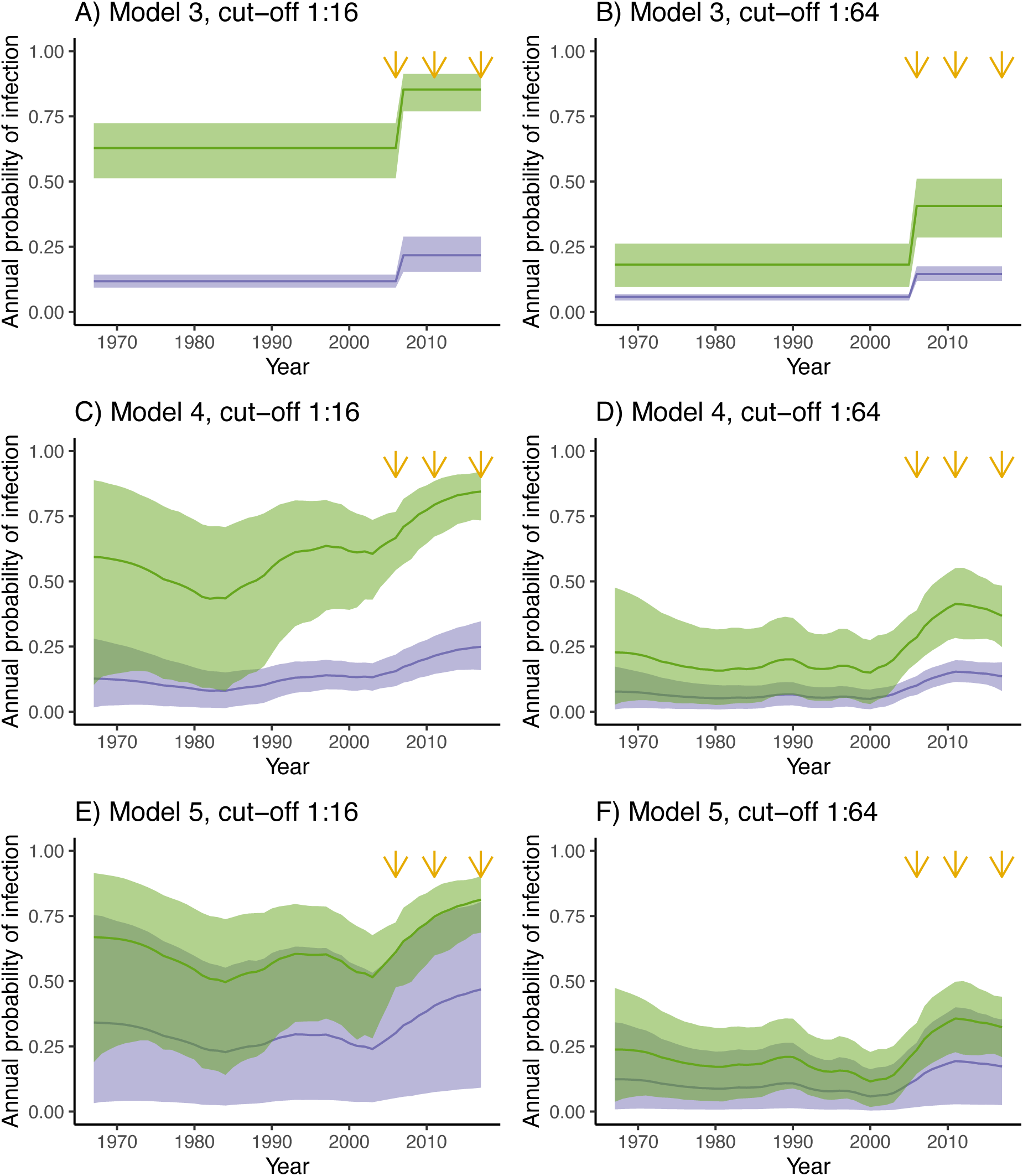
Estimated annual probability of infection. In green, the annual probability of infection for the 1-year-old age class and in purple for the rest of age classes. Results are presented for Model 3 (A, B), Model 4 (C, D) and Model 5 (E, F), and for two different seropositivity cut-offs, 1:16 (A, C, E) and 1:64 (B, D, F). Orange arrows indicate the years for which there is cross-sectional seroprevalence data.

**Figure 3.**
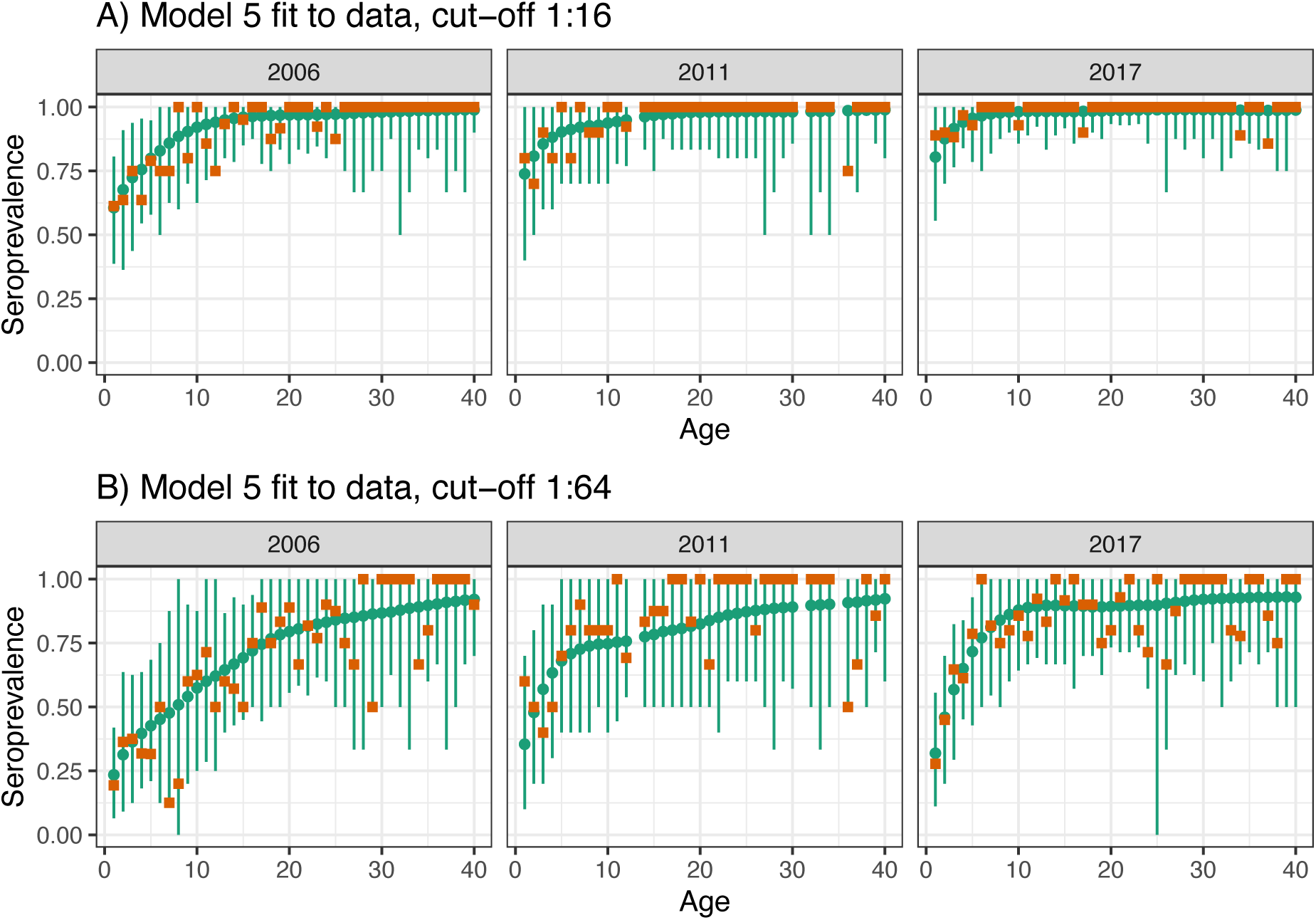
Model fit to data. Observed (brown squares) and modelled (green points) seroprevalence by age for the three cross-sectional serosurveys (2006, 2011 and 2017). Model fit is shown for the best model (Model 5) and for the two seropositivity cut-offs, (A) 1:16, and (B) 1:64. Green bars indicate the 95% binomial confidence intervals around the mean estimates, accounting for sample size. Similar results for Models 3 and 4 are shown in Figures S2 and S3 respectively.

Models 3 and 4 both estimated similar coefficients for the relative risk of infection at age 1 compared to other ages (median 7.89 and 6.56 for Models 3 and 4 respectively, for a seropositivity cut-off of 1:16, Table S1), which was lower for the more stringent seropositivity cut-off of 1:64 (median 3.32 and 3.17 for Models 3 and 4 respectively, Table S1). Model 5, which assumes an exponential decrease in the risk of infection through age, had a median estimate of the relative risk of infection at age 1 vs. age 2 of 3.32 and it increased up to 32.44 for age 1 vs. age 40 for a seropositivity cut-off of 1:16 (for a seropositivity cut-off of 1:64, these were 1.73 for age 1 vs. 2 and increased up to 7.93 for age 1 vs. 40).

Models 3, 4 and 5 provided a similarly good fit to the data, as indicated by the close estimates of the log-likelihood (Table 1). However, the best model according to the leave-one-out (LOO) information criterion, which accounts for over-parameterisation, was Model 5, closely followed by Model 4 (p=0.21 and p=0.13 for the seropositivity cut-offs of 1:16 and 1:64 respectively, i.e. we did not find a statistically significant difference in their predictive performance). Surprisingly, the step-function model (Model 3) ranked last, perhaps indicating that the LOO criterion is very sensitive to the abrupt change in FOI assumed in Model 3.

**Table 1.** Model comparison for the two different datasets, corresponding to two different seropositivity cut-offs. For each model comparison, the first row corresponds to the model with the largest expected log pointwise density (ELPD), which measures the model expected predictive accuracy. For each model, its log-likelihood and the difference in the Bayesian leave-one-out (LOO) estimate of the ELPD compared to the best model are shown.

| Model | log-likelihood | ΔLOO |
| --- | --- | --- |
| *Seropositivity cut-off 1:16* |  |  |
| 5 | -70.75 | 0.00 |
| 4 | -73.52 | -2.37 |
| 2 | -80.75 | -11.33 |
| 1 | -151.13 | -79.98 |
| 3 | -74.40 | -118.02 |
| *Seropositivity cut-off 1:64* |  |  |
| 5 | -148.95 | 0.00 |
| 4 | -152.35 | -3.05 |
| 2 | -153.96 | -5.16 |
| 1 | -214.20 | -63.06 |
| 3 | -149.33 | -82.41 |

We next used the FOI estimates from the two best models (Models 4 and 5) and data on the age structure of the population to reconstruct the overall EV-D68 seroprevalence in the population (Figure 4A) and the annual basic reproduction number (Figure 4B) for the period between the first and last cross-sectional serosurveys, 2006-2017. The two models estimate a progressive increase in the overall seroprevalence, driven by a progressive increase in the basic reproduction number between 2006 and 2017 (Figure 4). As expected, these estimates are higher when using the lower seropositivity cut-off (1:16).

**Figure 4.**
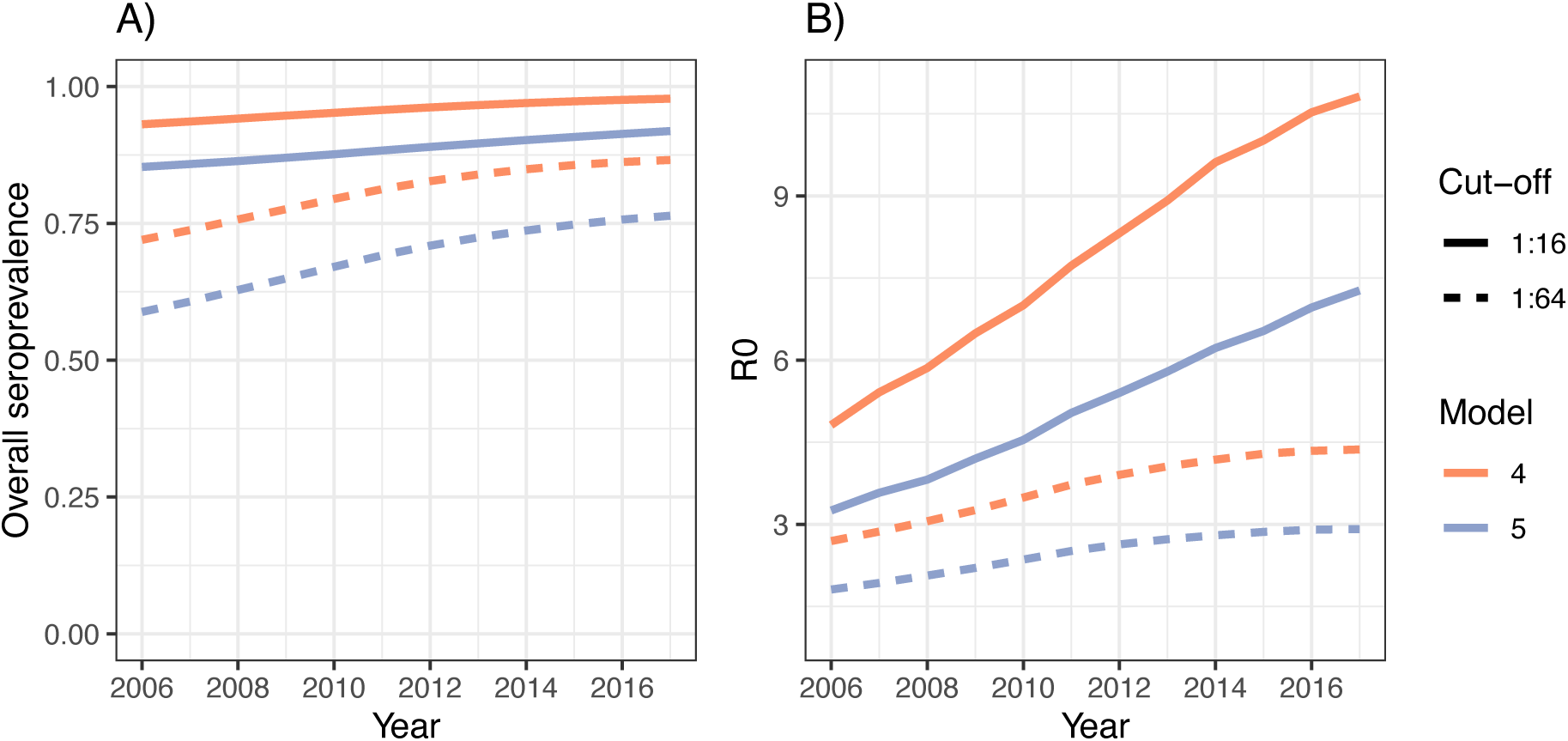
Seroprevalence and basic reproduction number. (A) Estimated overall (age-weighted) seroprevalence in the population and (B) basic reproduction number for the two best models (Models 4 and 5) and two different seropositivity cut-offs.

The amount of transmission or extent of virus circulation is better quantified by the number of infections than the FOI, which is sensitive to changes in the age structure of the population (e.g. driven by changes in birth rates) (18). Using the two best models (Models 4 and 5), we reconstructed the annual number of infections in each age class over time (Figures S9 and S10), which provided similar results. For a weak seropositivity cut-off of 1:16, the models predicted an increase in the number of infections in the 1-yo age group over time (Figures S9A and S10A), but a decrease over time in the subsequent age classes, with perhaps a slight increase in older age classes (Figure S9B and S10B). The decrease over time in infections in those aged 2, 3, etc. yo is due to the high and increasing seroprevalence in the 1-yo age class, which means only a small fraction of these children remain susceptible when they are 2, and so on. For the more stringent cut-off of 1:64, the model also predicts an increase over time in the number of infections in the 1-yo age group, that seems to plateau from around 2011 (Figure S9C and S10C). In this case, there is also a slight increase over time in the number of infections in the 2-and 3-yo, but a decrease in the older age classes (Figure S9D and S10D).

## Discussion

The model-based analysis of individual serological data for EV-D68 from three time points (2006, 2011 and 2017) in England presented here suggests an increase in transmission of EV-D68 that started during the 2000s. This coincides with the increased number of outbreaks of EV-D68-associated severe respiratory diseases reported worldwide since the late 2000s (2, 4).

These findings point to a clear increase in transmission as measured by the estimated annual probability of infection and the basic reproduction number (approximately, two-fold). However, the reconstructed annual number of new infections in each age class suggests that this increase is mostly driven by an increase in the total number of infections in children aged 12-23 months old. Increased transmission in the youngest age groups may be consistent with observed data showing higher and increasing numbers of respiratory illnesses associated with EV-D68 in these age groups (<5 yo) (19). However, these results on their own are unlikely to explain the worldwide emergence of AFM outbreaks reported since 2014. First, the analyses suggest that EV-D68 was already widely circulating before 2014. Second, AFM cases do not exclusively or predominantly occur in the youngest age groups. In the US, for example, AFM cases reported in 2014 had a median age of 7.1 yo (IQR, 4.8 – 12.1 yo) (20). In the UK, a study from 2018 reported 40 cases, of which only 22 were 0-5 yo (10). A study from European countries (2016) reported a median age of cases of 3.8 yo and a range of 1.6 – 9.0 yo. In Japan, a case series study of an AFM cluster reported an overall median age of cases of 4.4 yo (IQR, 2.6 – 7.7 yo) (21). Although susceptibility to EV-D68-related AFM may vary with age (22) (making it difficult to make a link between the inferred number of infections by age class and the observed number of cases by age), our model-based results suggest that incidence of infections in the age groups affected by AFM has not increased, despite the general increase in transmission. Therefore, the acquisition of or an increase in neuropathogenicity as well as transmission seems necessary to explain the emergence of AFM through an as-yet unidentified biological mechanism.

Our best-fitting models predict an increase in the number of infections in children 1-yo because of the high seroprevalence observed at age 1 (Figure 1) and also because we do not allow for re-infections. As a consequence, only a small number of infections occur at older ages (>1 yo), because most of the population has already been infected at age 1. Indeed, the three models that best explained the data required a higher FOI at age 1 (compared to the other age classes) to be able to explain the high seropositivity rates observed at this age. Although the extremely high seropositivity rates at age 1 are not generally found with other enteroviruses, they have been reported for EV-D68 in other places (23), e.g. the US (24), the Netherlands (17) and China (25, 26). Clarifying the origin and the meaning of this high prevalence of neutralising antibodies at this young age (the role of serum neutralising antibodies in protection against infection and diseases) should be a priority for EV-D68 research (23).

If serum neutralising antibodies are not a good correlate of protection against infection, the models may not capture well the age at which infections have increased, and it could be that the increase in infections is across age groups, not mainly in the 1 yo. Indeed, increased transmission may have also been associated with re-infections (and subsequent boosting of antibodies) in older age groups, consistent with the observed rise in geometric mean titers (GMTs) with age (15). However, it seems unlikely that secondary infections can lead to paralysis, based on data for poliovirus.

Whether antigenic escape can explain re-infections in older children is not fully understood. There is evidence of amino acid changes in the BC and DE loop regions of the VP1 (which are thought to be epitopes for neutralising antibodies) that might have resulted in altered antigenic properties (27, 28). However, although neutralisation assays conducted against different EV-D68 strains found some differences in neutralisation titers, study results have been inconsistent and their clinical and epidemiological significance unclear (15, 24).

Three major co-circulating EV-D68 clades (A-C) emerged globally in the 2000s (4) and have subsequently diversified, with only one monophyletic group (B1 and B3 genotypes) with a common ancestor in 2009 so far associated with AFM (29) (with the exception of one case associated with D1 in 2018 in France (30)). Individual viral lineages show rapid global spread, with recent outbreaks synchronised across Europe and the US representing circulation of the same dominant genotypes (e.g. co-circulating B3 and D1 in 2018). In vitro studies of the neurotropism of these viruses compared with the ancestral strains have yielded conflicting results as to whether neurotropism has increased (22, 31, 32). The timing of the increase in transmission estimated here based on the analysis of the serology data roughly corresponds to the genetic emergence of clade B around 2007, and thus one could hypothesise that increased virus transmissibility is a trait associated with this clade. More efficient viral replication may enhance transmission as well as the probability of virus reaching the central nervous system, although changes in receptor usage could also play a role.

Our ability to recover more complex changes in transmission is limited by the data available. It would not be surprising if EV-D68 has exhibited biennial (or longer) cycles of transmission in England over the last few years, as it has been shown in the US (7) and is common for other enteroviruses (33). However, it is difficult to recover changes at this finer time scale with serology data unless sampling is very frequent (at least annual). Therefore, our study can only reveal broader long-term secular changes.

This work shows the value of modelling age-stratified seroprevalence data from consecutive cross-sectional studies in the understanding of the epidemiology of diseases caused by emerging human enteroviruses. The dynamics of most enterovirus serotypes over relatively long time scales have been shown to be driven by population immunity (33). However, in rare instances, enterovirus serotypes have emerged as important causes of diseases after many years of circulation causing diseases at a much lower rate or even silently circulating. For example, coxsackievirus A6 (CVA6) has emerged as the main serotype causing HFMD worldwide over the last decade (34). Analysis of CVA6 serological data could help confirm whether there have been changes in transmission contributing to this emergence. Finally, this work also shows the need to better understand and interpret individual serological data in terms of previous exposure and protection against infection and disease. This would help refining analytical approaches such as those used here to infer population-level processes.

## Materials and Methods

### Serological data

We use data from three retrospective cross-sectional studies analysing serum samples representative of England’s population in 2006 (n=516), 2011 (n=504) and 2017 (n=566) and available through the National Seroepidemiology Programme at Public Health England (35). The number of samples per age class and cross-sectional study, for the age classes used in the analyses are shown in Table S2. The neutralisation assay method and results from serological testing of the 2006 and 2017 sample sets have been previously described in (15). Neutralisation assays measured neutralising antibody titers against a B3 strain (15, 29), but Kamau et al. showed similar neutralisation effects across three different EV-D68 strains (15).

### Statistical analysis

The FOI is the rate at which seronegative (susceptible) individuals become seropositive (infected). Cross-sectional age-stratified seroprevalence data can be used to estimate the FOI through so-called catalytic models (36). Catalytic models avoid modelling the dynamics of infected individuals directly; rather, they assume an unspecified mechanism that results in a susceptible individual becoming infected, its magnitude defining the FOI. For seroprevalence data, these models rely on the idea that the age and serostatus of an individual provide information on the probability of infection for the years between birth and the serosurvey.

General approach. Let *S*^*τ*^ (*t*) be the proportion of susceptible (seronegative) individuals born at time *τ* at some subsequent time *t* ≥ *τ*. We assume that after infection (seroconversion), individuals cannot become susceptible again (i.e. we do not include seroreversion). This seems reasonable since seroprevalence does not show a decline through age (Figure 1). This results in the following equation:

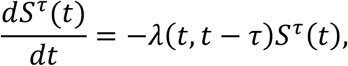

where we suppose that the FOI, *λ* (*t, t*− *τ*) > 0, can vary over time *t*and with the age of an individual *a* =*t* − *τ*.

To avoid the effect of maternal antibodies, we exclude individuals <1 year of age from the analysis and we assume that individuals enter the model at age 1 being seronegative, so that *S* ^*τ*^ (*τ* + 1) = 1. This permits the solution:

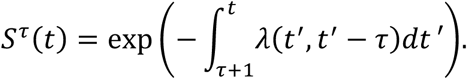

We assume that the historical FOI is different each year but constant within it. This is important for enteroviruses, as most serotypes do not circulate every year, but rather in longer cycles of two or more years (33). This means we can rewrite the above equation as:

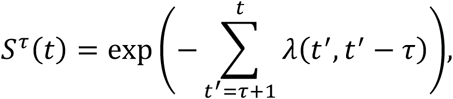

where *t* is now a discrete measure of time in years.

An individual is deemed “seropositive” if their antibody titer exceeds a given cut-off. We let *X* ^*τ*^ (*t*|*c*) denote the serostatus (seropositive or seronegative) of an individual born in year *τ* in a subsequent year *t*≥ *τ* assuming a seropositivity cut-off *c*. Then the probability an individual is seropositive in year *t*is given by:

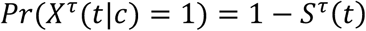

which implicitly assumes that antibody titers are long lasting, and as such, once an individual has seroconverted there is no waning immunity.

We further assume that all individuals born in the same year have been exposed to a common historical FOI throughout their lives. We also assume that testing uncovers seropositivity with 100% accuracy. The count of seropositive individuals within a serosurvey in year *t* can then be modelled using a binomial distribution:

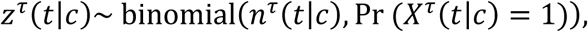

where *n* ^*τ*^ (*t*|*c*) indicates the sample size in year *t*for individuals born in year *τ* as collected during the serosurvey; and 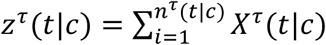 is the count of those individuals who are seropositive.

We test five different models representing different hypotheses about how the FOI changes over time *t* (in years), and also through age *a*. The mathematical details describing the different models are given below.

Model 1: Constant FOI over time and age,

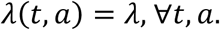

This model has a single parameter, *λ*.

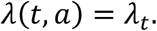

Model 2: Different and independent FOIs each year but FOI is age-independent,

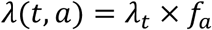

Model 3: Step-function model with increased FOI at age 1,

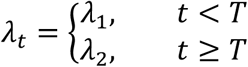

where

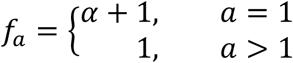

and

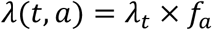

The parameter *T*, the year when the force of infection changes, is estimated, and the parameter *α* > 0, describing the increase in FOI at age 1 compared to the other ages, is also estimated.

Model 4: Random walk model of order one, with increased FOI at age 1,

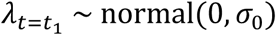

where

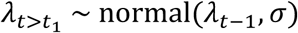

and

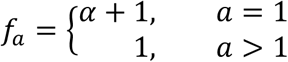

Model 5: Random walk model of order one, with increased FOI at age 1 and exponential decrease in FOI through age,

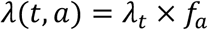

where

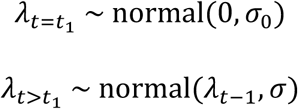

and

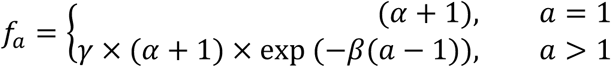

This model is an extension of Model 4.

The annual probability of infection, which is the proportion of the susceptible population that will become seropositive in a given year, can be derived from the FOI. With our approach, the annual probability of infection for year *t* and age *a, p*(*t, a*), is *p*(*t, a*) =1 − exp(−*λ*(*t, a*)).

Because seroprevalence in adults reaches almost 100% from about the 20-yo age class for a seropositivity cut-off of 1:16, and from the 30-yo for a more stringent cut-off of 1:64 (Figure 1), we fit the models to the data for the first N=40 age classes.

The models were implemented in RStan (37) and fitted to the data using MCMC. Four independent chains were simulated, each of 10,000 iterations, with a warmup of 3,000. Convergence was checked using the Rhat function.

### Annual overall seroprevalence, basic reproduction number and reconstructed number of infections in each age class

Using the estimates of the FOI from the catalytic models above and data on population structure, we can estimate the overall (age-weighted) seroprevalence in the population each year *t*. We used data on the population structure in England for the years 1998, 2008 and 2018 (Figure S8), from (38). The size of each age class for the years in between was obtained by linear interpolation.

We then used age-specific estimates of the FOI and seroprevalence to obtain annual estimates of the basic reproduction number, using equation (3) from Farrington et al. (39), which gives the relationship:

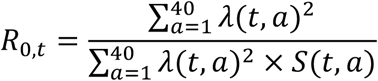

where *S*(*t, a*) is the proportion seronegative at age *a* and year *t*. Note that we further make the hypothesis that age-specific mortality rate is zero in the 40 age classes that we consider.

Finally, we reconstructed the annual number of (new) infections in each age class using the estimates of the FOI and population structure data. To reconstruct the number of infections in a given year *t* and age class *a*, we first reconstructed the proportion seronegative in the age class *a-1* until year *t-1*, and then derived the proportion who would seroconvert during year *t*. We then multiplied that proportion by the population size of the corresponding age class and year.

## Data Availability

All data produced in the present study are available upon reasonable request to the authors.

## Acknowledgements

M.P.-S. is a Sir Henry Dale Fellow jointly funded by the Wellcome Trust and the Royal Society (grant number 216427/Z/19/Z). M.P.-S. and N.C.G. acknowledge funding from the MRC Centre for Global Infectious Disease Analysis (reference MR/R015600/1), jointly funded by the UK Medical Research Council (MRC) and the UK Foreign, Commonwealth & Development Office (FCDO), under the MRC/FCDO Concordat agreement and is also part of the EDCTP2 programme supported by the European Union. Work in P.S.’s lab was supported by a Wellcome ISSF grant (ISSF204826/Z/16/Z). We thank the PHE Sero-Epidemiology Unit for access to residual samples for this public health investigation.

## Supplementary Material

**Figure S1.**
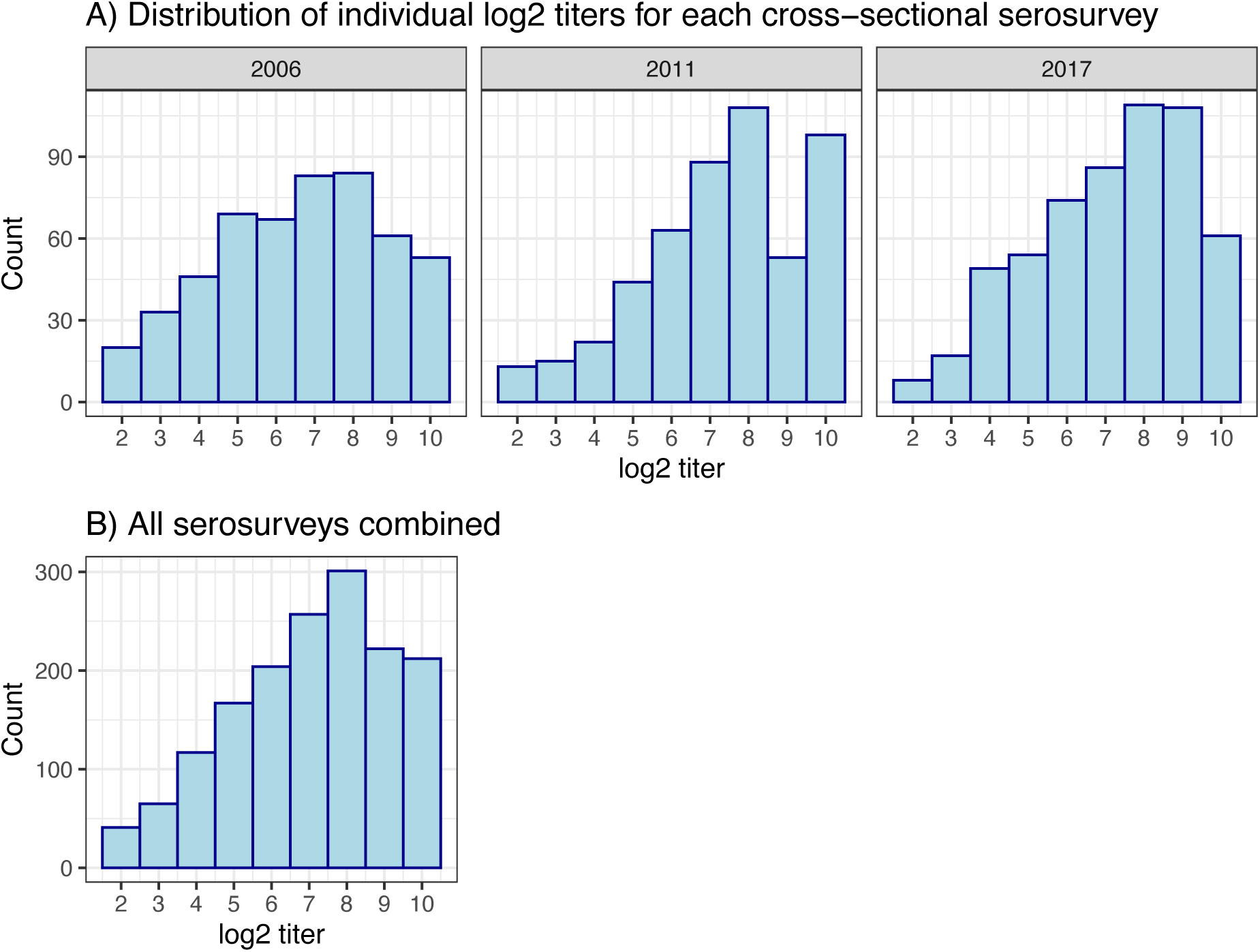
Individual log2 titer distributions. (A) For each serosurvey. (B) For all surveys combined.

**Figure S2.**
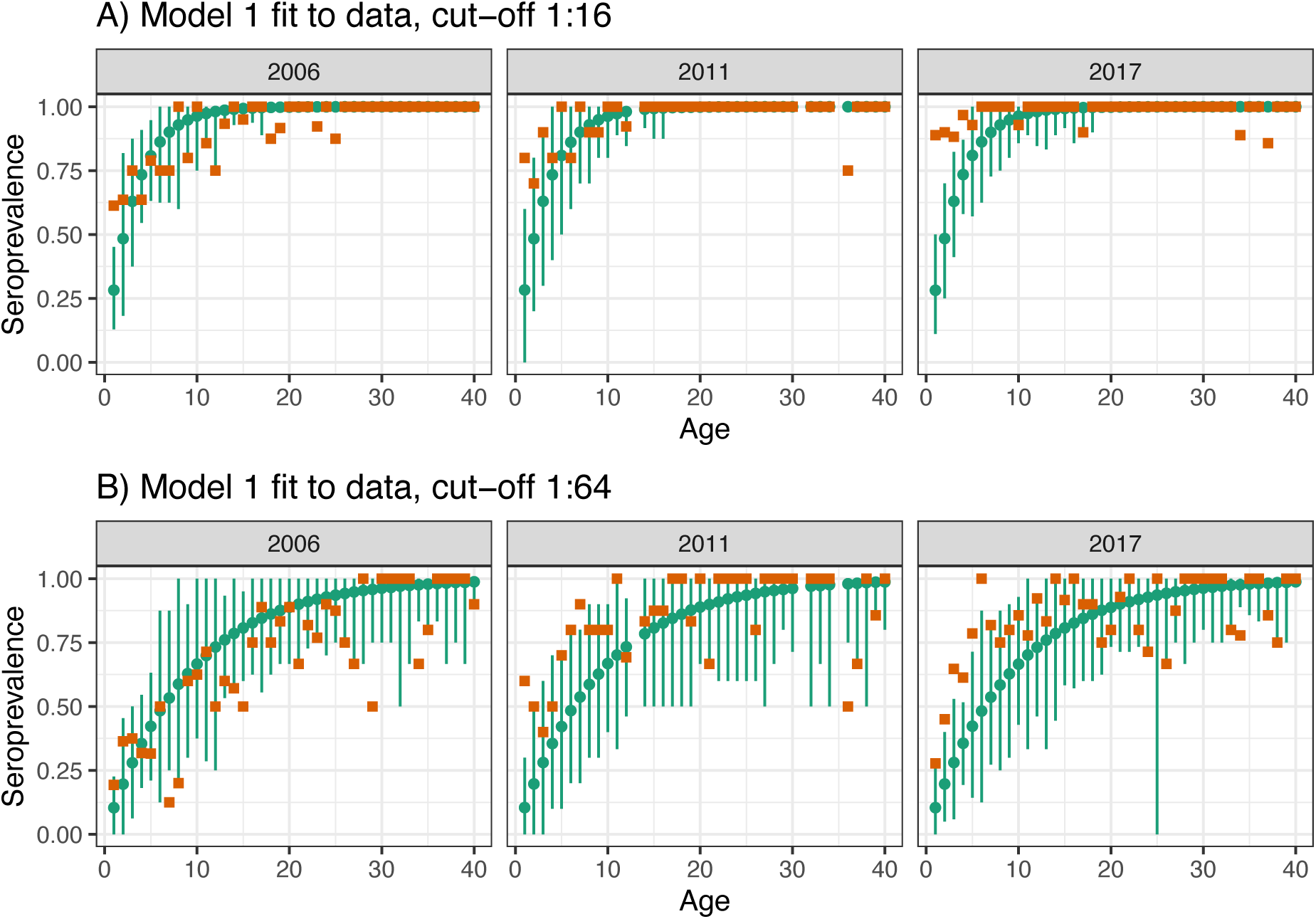
Model 1 fit to data. Observed (brown squares) and modelled (green points) seroprevalence by age for the three cross-sectional serosurveys (2006, 2011 and 2017). Model fit is shown for Model 1 and for the two seropositivity cut-offs, A) 1:16, and B) 1:64. Green bars indicate the 95% binomial confidence intervals around the mean estimates, accounting for sample size.

**Figure S3.**
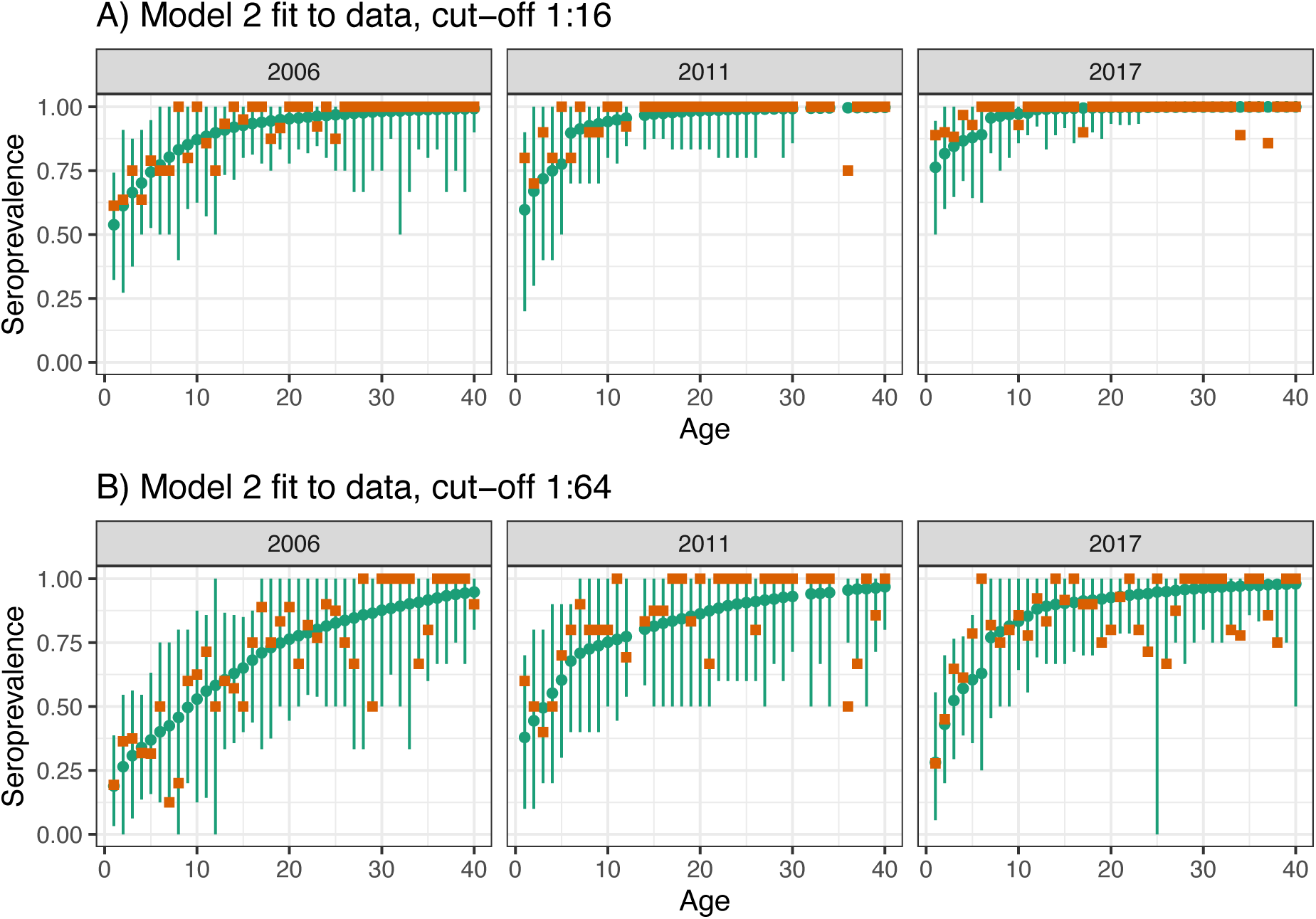
Model 2 fit to data. Observed (brown squares) and modelled (green points) seroprevalence by age for the three cross-sectional serosurveys (2006, 2011 and 2017). Model fit is shown for Model 2 and for the two seropositivity cut-offs, A) 1:16, and B) 1:64. Green bars indicate the 95% binomial confidence intervals around the mean estimates, accounting for sample size.

**Figure S4.**
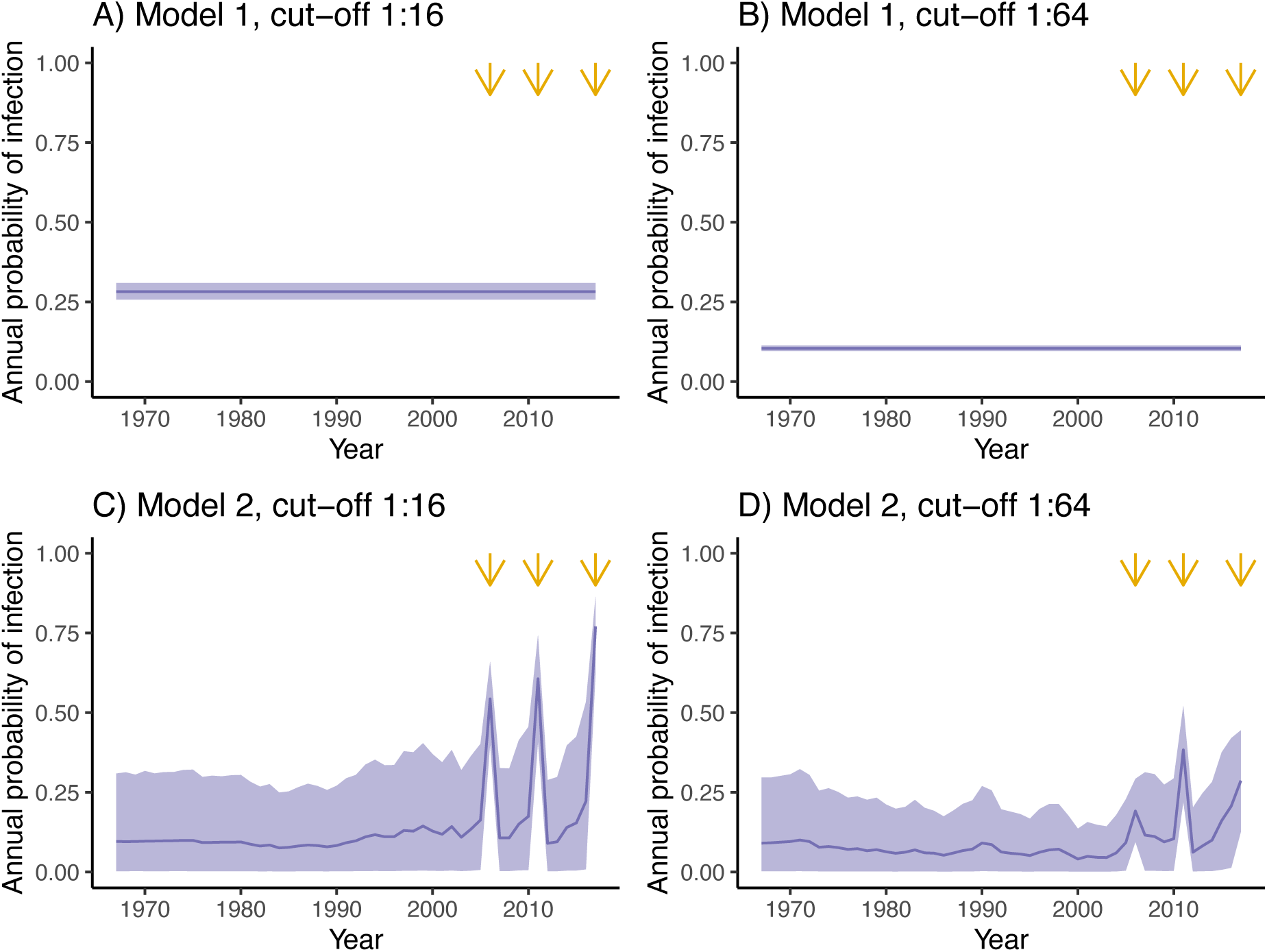
Estimated annual probability of infection for Models 1 and 2. Results are presented for Model 1 (A, B) and Model 2 (C, D), and for two different seropositivity cut-offs, 1:16 (A, C) and 1:64 (B, D). Orange arrows indicate the years for which there is cross-sectional seroprevalence data.

**Figure S5.**
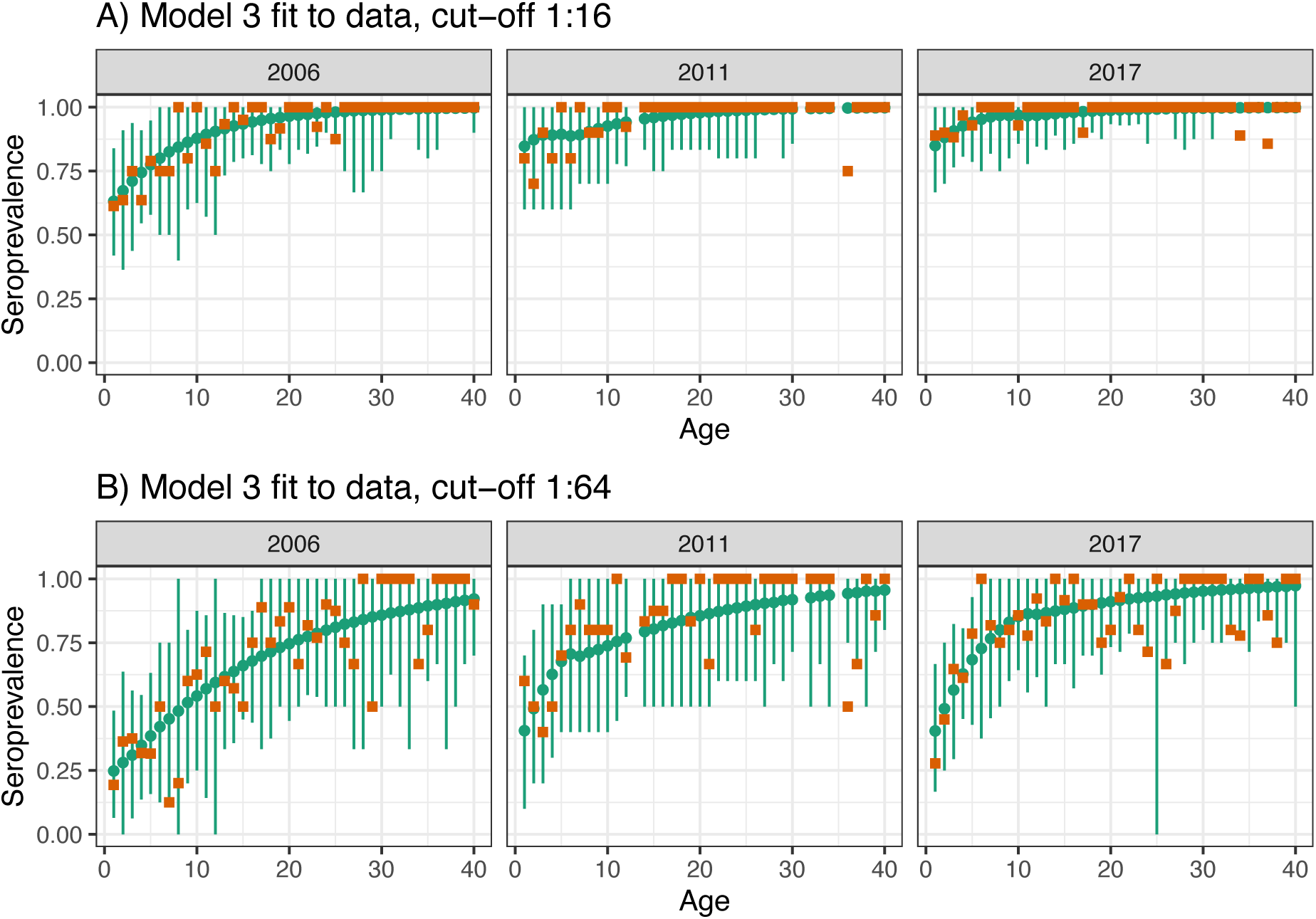
Model 3 fit to data. Observed (brown squares) and modelled (green points) seroprevalence by age for the three cross-sectional serosurveys (2006, 2011 and 2017). Model fit is shown for Model 3 and for the two seropositivity cut-offs, A) 1:16, and B) 1:64. Green bars indicate the 95% binomial confidence intervals around the mean estimates, accounting for sample size.

**Figure S6.**
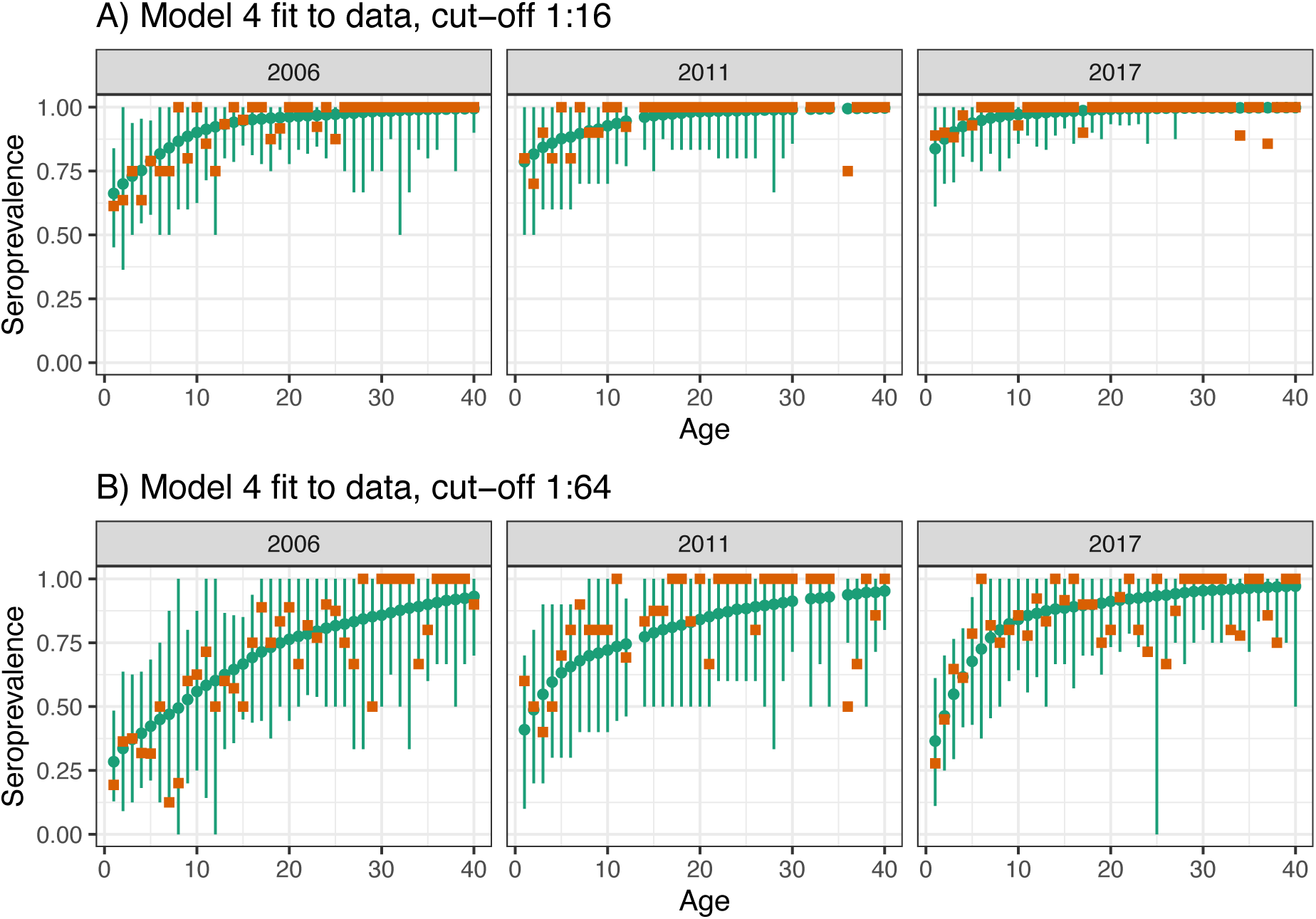
Model 4 fit to data. Observed (brown squares) and modelled (green points) seroprevalence by age for the three cross-sectional serosurveys (2006, 2011 and 2017). Model fit is shown for Model 4 and for the two seropositivity cut-offs, A) 1:16, and B) 1:64. Green bars indicate the 95% binomial confidence intervals around the mean estimates, accounting for sample size.

**Figure S7.**
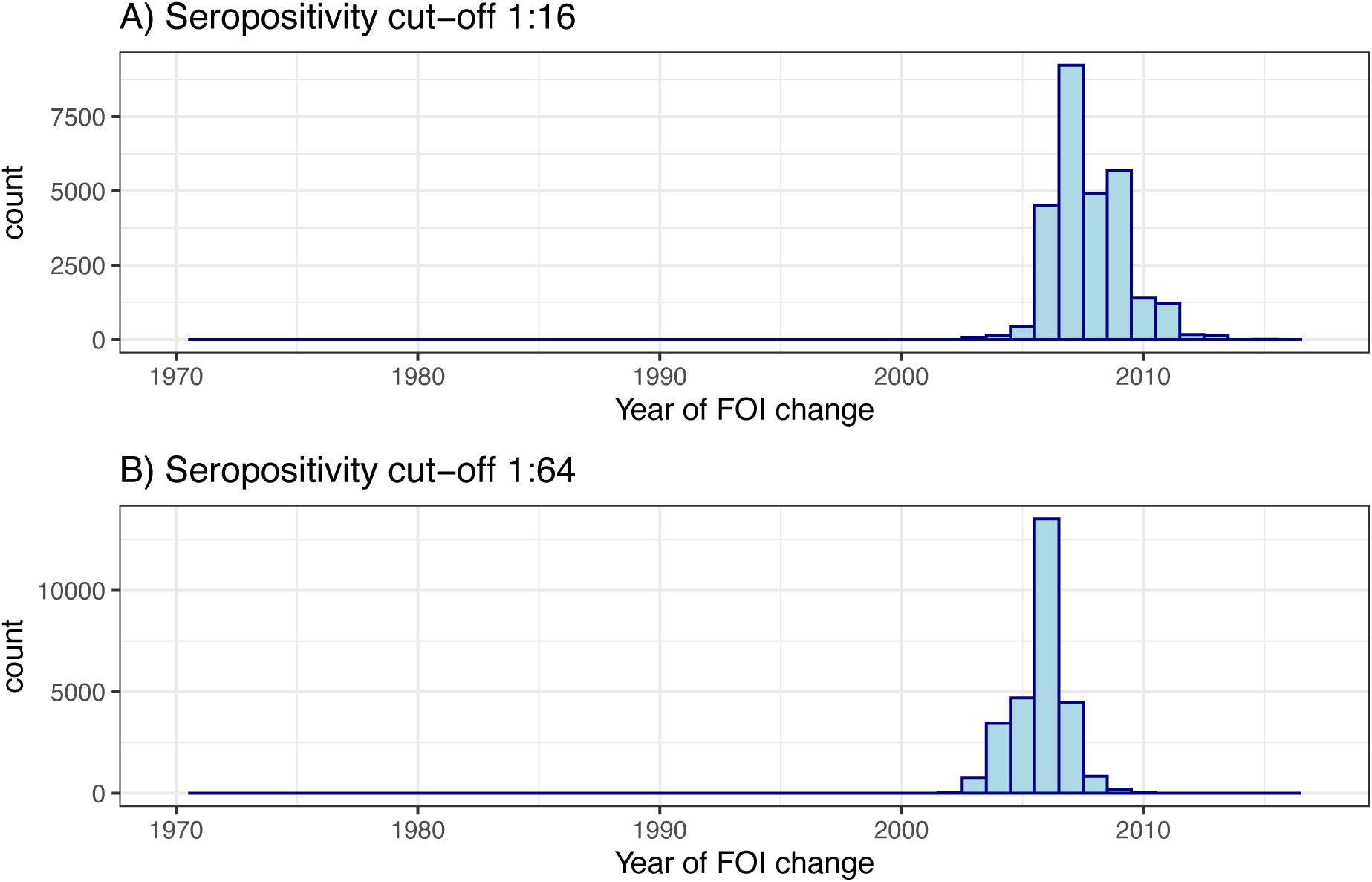
Posterior distribution for the estimated year of the change in FOI for the step-function model (Model 3) for seropositivity cut-offs of (A) 1:16, and (B) 1:64.

**Figure S8.**
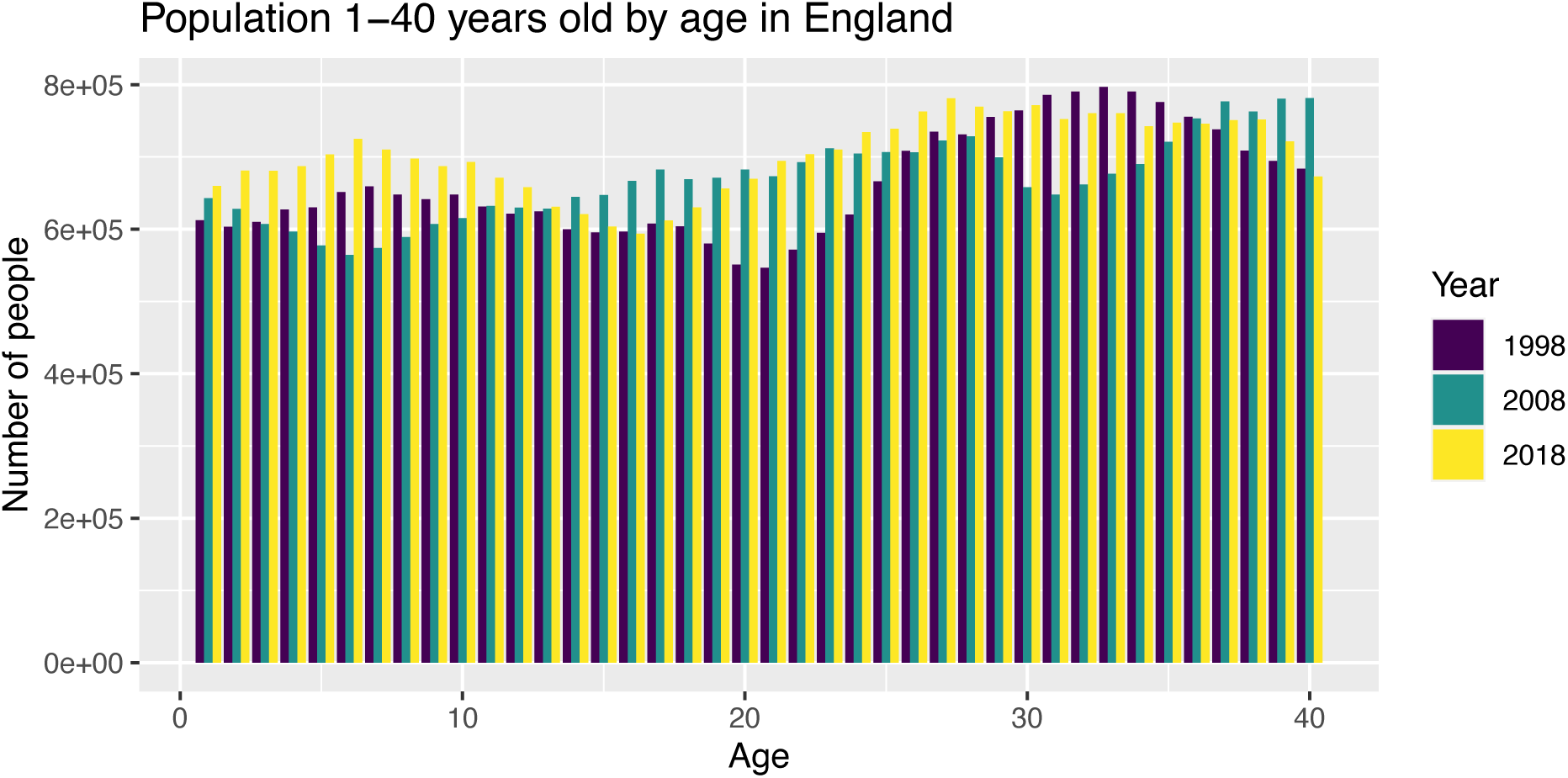
England population age structure for the years 1998, 2008 and 2018.

**Figure S9.**
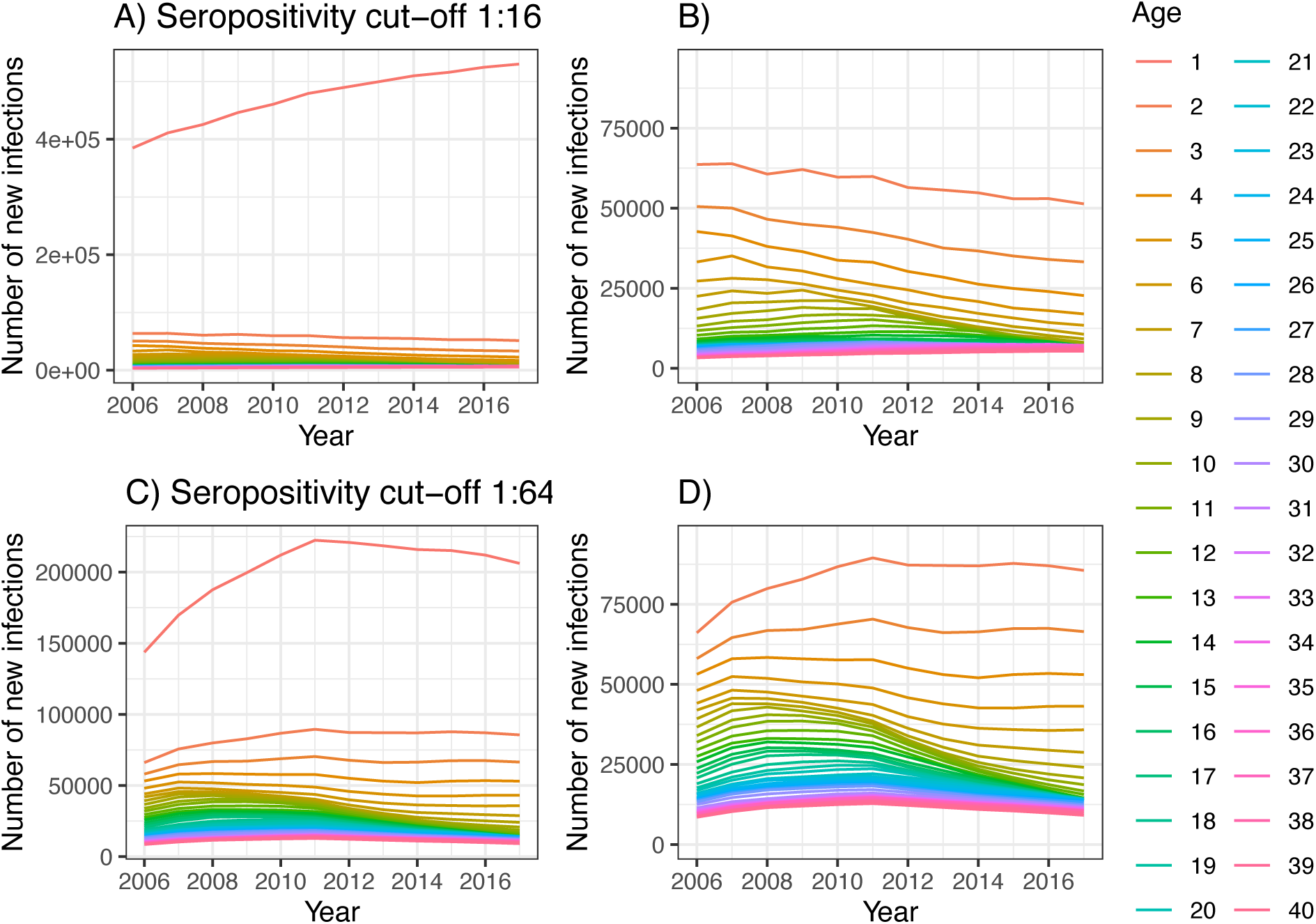
Reconstructed number of infections with Model 5. Reconstructed annual number of new infections in each age class using the best model (Model 5) for the two seropositivity cut-offs: 1:16 (A, B) and 1:64 (C, D). Note that (B) and (D) are a zoom in of (A) and (C) respectively for the age classes above 1 year of age.

**Figure S10.**
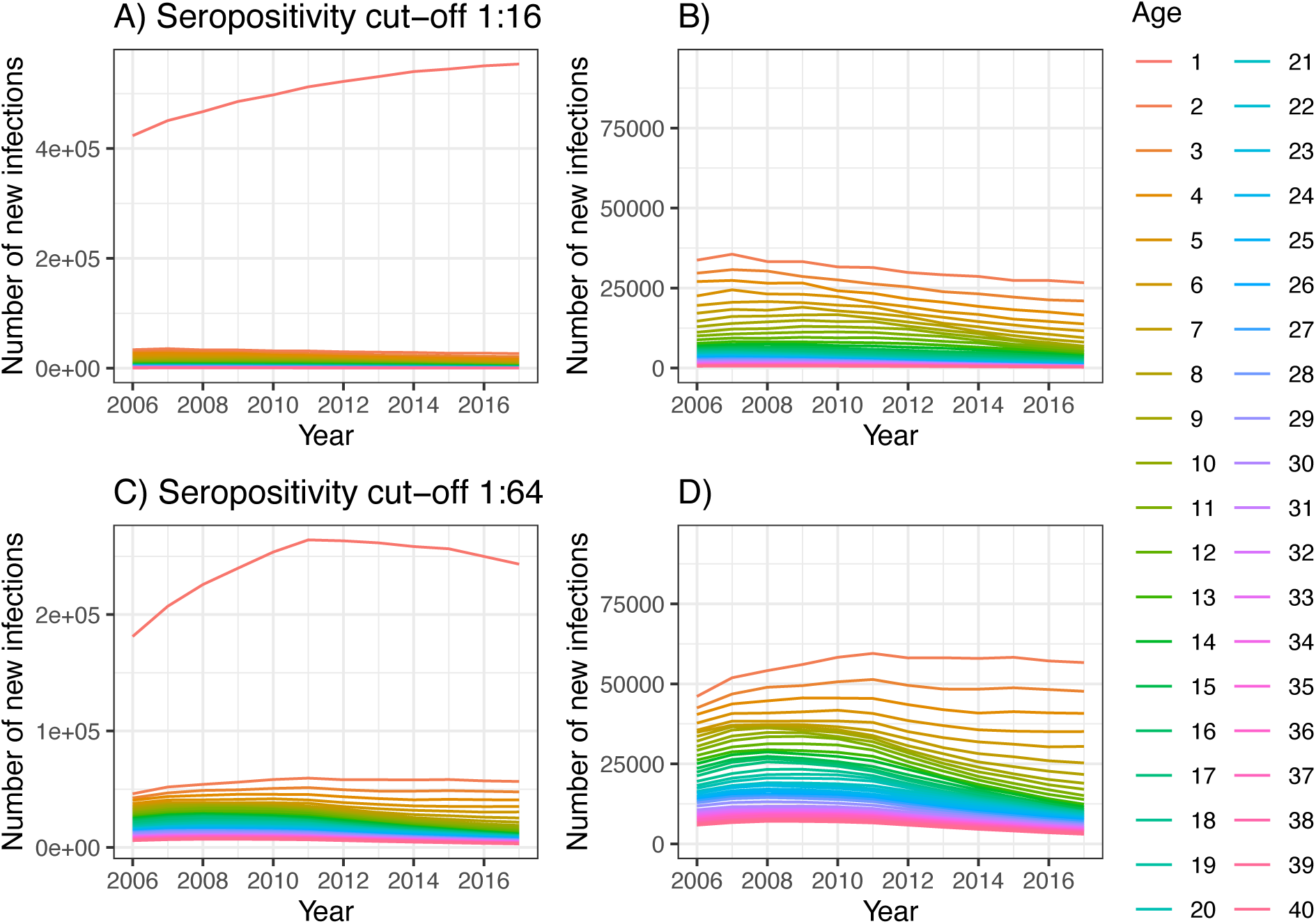
Reconstructed number of infections with Model 4. Reconstructed annual number of new infections in each age class using the second best model (Model 4) for the two seropositivity cut-offs: 1:16 (A, B) and 1:64 (C, D). Note that (B) and (D) are a zoom in of (A) and (C) respectively for the age classes above 1 year of age.

**Table S1.** Model parameter estimates. Estimates of the parameters describing the age-related increased risk of infection in the 1-year-old age class (**α**) and the other age classes (**Ψ**,**α**).

| Parameter | Model 3 | Model 4 | Model 5 |
| --- | --- | --- | --- |
| ***Seropositivity cut-off 1:16*** |  |  |  |
| $\alpha$ | 6.89 (4.18,11.05) | 5.56 (3.05, 9.97) | 2.10 (0.07, 15.85) |
| $\gamma$ | - | - | 0.32 (0.16, 0.58) |
| $\beta$ | - | - | 0.06 (0.02, 0.13) |
| ***Seropositivity cut-off 1:64*** |  |  |  |
| $\alpha$ | 2.32 (0.95, 4.14) | 2.17 (0.97, 4.00) | 1.14 (0.03, 13.42) |
| $\gamma$ | - | - | 0.60 (0.31, 1.06) |
| $\beta$ | - | - | 0.04 (0.01, 0.07) |

**Table S2.** Number of samples in each age class for the age classes used in the analyses.**40** 10 5 2

| Age class | 2006 | 2011 | 2017 |
| --- | --- | --- | --- |
| 1 | 31 | 10 | 18 |
| 2 | 11 | 10 | 20 |
| 3 | 16 | 10 | 17 |
| 4 | 22 | 10 | 31 |
| 5 | 19 | 10 | 14 |
| 6 | 8 | 10 | 8 |
| 7 | 8 | 10 | 11 |
| 8 | 5 | 10 | 8 |
| 9 | 10 | 10 | 10 |
| 10 | 8 | 10 | 14 |
| 11 | 7 | 9 | 9 |
| 12 | 4 | 13 | 13 |
| 13 | 15 | 0 | 6 |
| 14 | 14 | 12 | 9 |
| 15 | 20 | 8 | 12 |
| 16 | 16 | 8 | 7 |
| 17 | 9 | 6 | 10 |
| 18 | 8 | 6 | 10 |
| 19 | 12 | 6 | 8 |
| 20 | 9 | 6 | 15 |
| 21 | 12 | 6 | 14 |
| 22 | 11 | 5 | 14 |
| 23 | 13 | 5 | 15 |
| 24 | 10 | 5 | 7 |
| 25 | 8 | 5 | 1 |
| 26 | 4 | 5 | 3 |
| 27 | 3 | 2 | 8 |
| 28 | 3 | 3 | 6 |
| 29 | 4 | 5 | 8 |
| 30 | 4 | 7 | 5 |
| 31 | 8 | 0 | 8 |
| 32 | 2 | 2 | 4 |
| 33 | 3 | 3 | 5 |
| 34 | 6 | 2 | 9 |
| 35 | 5 | 0 | 7 |
| 36 | 6 | 4 | 6 |
| 37 | 3 | 3 | 7 |
| 38 | 4 | 2 | 4 |
| 39 | 3 | 7 | 4 |
| **40** | 10 | 5 | 2 |

